# Anemia and associated factors among 6-59 months age children attending health facilities in Northeast Ethiopia: A facility based cross sectional study

**DOI:** 10.1101/2022.09.30.22280545

**Authors:** Wubshet Fentaw, Tefera Belachew, Assefa Andargie

## Abstract

**Background:** In the early stages of life, anemia leads to severe negative consequences on the cognitive, growth and development of children. The Ethiopian demographic and health survey showed an increasing trend of anemia nationally.

**Objective:** The aim of this study is to assess the magnitude and factors associated with anemia among under-five children.

**Methods:** A health facility based cross sectional study was conducted among 409 systematically selected children 6-59 months attending services at public and private health institutions in Kombolcha Town, Northeast Ethiopia. Data were collected using structured questioner from mother or caretakers. The data entry and analysis were done by SPSS version 20. Binary logistic regression was fitted to determine associations. The odds ratio with the 95% confidence intervals were reported.

**Results:** From the total participant 213(53.9%) were males with the mean age of 26 months (SD±15.2). The overall prevalence of anemia was 52.2% (95% CI, 46.8%-57%). Being in the age of 6-11 months (AOR= 6.23, 95% CI: 2.44, 15.95), 12-23 months (AOR= 3.74, 95%CI: 1.63, 8.60), maternal age ≥30 years (AOR=0.37 (0.18, 0.77), exclusive breast feeding until six months (AOR=0.27, 95% CI: 0.16, 0.45), having low dietary diversity score (AOR=2.61, 95% CI: 1.55, 4.38), having history of diarrhea (AOR= 1.87, 95% CI: 1.12, 3.12) and having the lowest family monthly income (AOR=16.97, 95% CI: 4.95, 58.20) were identified as factors associated with anemia.

**Conclusion:** The magnitude of anemia in children was a public health problem in the study area. child age, maternal age, exclusive breastfeeding, dietary diversity score, diarrhea and family income were found to be associated with anemia.

## Introduction

Anemia is a major health problem throughout the world. The magnitude of the problem in developing countries is high since they were more exposed to various health and socioeconomic problems, which are directly or in directly related with anemia [1]. Though all age groups can develop anemia due to various factors, under-five children are among the most vulnerable age groups [2]. According to the World Health organization (WHO) 2015 report 43% of under five children were anemic, with regional variations of 62.3% in African, 53.8% in South-East Asia and 21.9% in Western Pacific Region [3]. In East Africa, approximately 75 % of under-five children have anemia with the prevalence’s ranging between 44 and 76 % [4]. It was estimated that 57% of children under five years in Ethiopia had anemia [5].

Anemia among under five-children is a condition, which has serious consequences including growth retardation, poor immune system and increased susceptibility to diseases and death and has severe socio-economic consequences for families and communities [6–9]. It creates long-term effect among female children resulting in low-birth-weight babies and postpartum hemorrhage especially young children from low-income families have a higher risk for developing iron deficiency anemia [10].

In developing countries, insufficient dietary iron is considered as the primary cause of anemia in children [7,9,10]. Other factors like parasitic infections, maternal age, maternal educational status, maternal marital status, Ante Natal Care (ANC) visit, child’s place of birth, frequency of complementary feeding per day, loss of appetite, breastfeeding status, and family monthly income have significant effect in the differentials of anemia occurrence [5,11,20,12–19].

Several studies has been conducted across different parts of Ethiopia on childhood anemia [12,13,15,17,20–23]. However, there is scarcity of evidence on childhood anemia in the Northeastern part of the country where drought and food insecurity are rampant in the area. Therefore, this study was conducted to determine the prevalence and factors associated with anemia among under five children visiting health facilities in Kombolcha town, located in the Northeastern part of the country under the Amhara regional state.

## Materials and methods

### Study design and population

This study employed a facility based cross-sectional design in Northeast Ethiopia (Kombolcha town). The study population was all under-five years children who visited selected public and private health facilities for any services in the town during the study period.

### Study Area and period

The study was conducted in Kombolcha town located in the North-eastern part of Ethiopia in Amhara Regional State, South Wollo zone at a distance of 378 km from the capital Addis Ababa to the Northeast. The town is an important commercial and industrial center in the northeast Ethiopia. The study was conducted from February 1 to April 30, 2020.

### Eligibility criteria

All children who have been living above 6 months in the area and their age 6 to 59 months who visited the selected health institutions were included in this study. Whereas, children with known bleeding disorder, severe illness, mentally ill mother (care taker), and children who have taken blood and blood products (blood transfused) for the last 3 months were excluded from the study.

### Sample size and sampling procedure

The sample size was determined based on the single population assumption taking the prevalence of anemia to be 41.1% with 5 % margin of error and 95% confidence level [12]. By considering 10% non-response rate the final sample size was estimated to be 409. The list of health institutions was obtained from the town administration health office and were stratified into public and private. Then, half of the public and one-third of the private institutions were selected using simple random sampling technique (lottery method). The study subjects were selected using systematic sampling method.

### Outcome measurement

**Anemia**, in under five children, was defined as hemoglobin level below 11g/dl. In this study, the Hgb level was adjusted to altitude. Since the study area is found 1842m above sea level, 0.8 g/dl was added to the standard Hgb cut-off value. Therefore, children with Hgb of 11.8g/dl or more were considered as normal while children who had Hgb value *<*11.8 g/dl were considered as anemic [5].

### Data collection procedures

Pretested and structured questionnaire was used to collect socioeconomic and demographic characteristics of the family, breast feeding status and dietary diversity of the child and other risk factors by interviewing mother/caregivers of the child. The questionnaire was adapted from previous similar literatures and prepared in English, translated in to the local language, and then back to English to ensure its consistency. Six trained nurses and six laboratory technicians conducted the data collection.

### Blood specimen collection and examination

After asking the willingness of parents/care givers, a capillary blood samples were collected from children under strict aseptic precautions by the trained laboratory technicians. The hemoglobin concentration was measured using HemoCue Hb 201 analyzer.

### Stool specimen collection and examination

A clean plastic container marked with an identification number was used to collect about 2 mg of stool sample from each child. Then, stool wet mount smear was prepared using saline and/or iodine solution for direct microscopic identification of intestinal parasites within 30 minutes of sample collection. The direct smear was examined first by 10x and then 40x objective for detection of helminths eggs, larvae and protozoan parasites by experienced medical laboratory technicians.

### Data quality control

The quality of data was assured through careful designing, translation, pre-testing of the questionnaire and training of data collectors. Every day after data collection, the filled questionnaire was checked for completeness by the principal investigator, supervisors, and correction made accordingly for the next day. Standard operating procedures (SOPs) and manufacturers’ instruction was strictly followed for all laboratory activities. All laboratory reagents were checked for their expiry date. Samples were checked whether they are in the acceptable criteria like-volume, consistency and collection time. Microscopic slides and cover glasses were checked for cleanliness.

### Data processing and analysis

The collected data were entered in to Epidata version 3.1 and exported to SPSS version 26.0 for analysis. Descriptive statistics like frequencies, proportions and summary measures were computed. Binary logistic regression was employed to identify factors associated with anemia. First bivariate binary logistic regression was performed and variables found to be significant at P-value <0.25 were imputed into multiple logistic regression. A p-value *<* 0.05 was used to indicate statistical significance in the multiple logistic regression model. Both the crude and adjusted Odds Ratio were reported as effect measures. The results were presented using tables, figures and text narratives.

## Results

### Characteristics of study participants

Out of 409 participants 395 participants responded correctly making the response rate of 96.6%. The rest participants were refusals. About 213(53.9%) were males with the mean age of 26 months (SD±15.2). From the total of mothers interviewed, 211 (53.4%) were attended secondary education and above; and about 330 (83.5%) were from urban residence. Regarding the marital status of mothers, 357 (90.3%) were married (Table 1).

**Table 1:** Socio-demographic characteristics of children under-five years attending health institutions of Kombolcha town, Northeast Ethiopia (n=395).

| **Variables** | **Category** | **Frequencies** | **Percent** |
| --- | --- | --- | --- |
| Child Sex | Male | 213 | 53.9 |
|  | Female | 182 | 46.1 |
| Child age | 6-11 | 72 | 18.2 |
|  | 12-23 | 128 | 32.4 |
|  | 24-35 | 80 | 20.3 |
|  | 36-47 | 57 | 14.4 |
|  | 48-59 | 58 | 14.7 |
| Residence | Urban | 330 | 83.5 |
|  | Rural | 65 | 16.5 |
| Religion | Orthodox | 124 | 31.4 |
|  | Muslim | 265 | 67.1 |
|  | Protestant | 6 | 1.5 |
| Maternal age | 18-27 | 219 | 55.4 |
|  | 28-38 | 168 | 42 |
|  | >39 | 8 | 2 |
| Marital status | Single | 29 | 7.3 |
|  | Married | 357 | 90.4 |
|  | Divorced | 7 | 1.8 |
| Ethnicity | Amhara | 374 | 94.7 |
|  | Tigre | 17 | 4.3 |
|  | Other | 4 | 1 |
| Educational status of mothers | No formal education | 37 | 9.4 |
|  | Primary education | 147 | 37.2 |
|  | Secondary and above | 211 | 53.4 |
| Mothers' occupation | House wife | 304 | 77 |
|  | Employed | 81 | 20.5 |
|  | Student | 9 | 2.3 |
| Monthly family income (ETB) | <1500 | 62 | 15.7 |
|  | 1501-3000 | 67 | 17 |
|  | 3001-4500 | 201 | 50.9 |
|  | >4500 | 65 | 16.5 |
| Family size | <4 | 158 | 40 |
|  | 4 and above | 237 | 60 |

143 **Abbreviations:** ETB, Ethiopian Birr

### Nutrition and disease morbidities

Out of 395 study subjects, 171 (43.3%) had diarrhea from which 102 (49.8%) were anemic. Most of 256 (64.2%) of children got adequate dietary diversity score (four and more food groups) per 24 hours out of them 104 (40.6) were anemic (Table 2).

**Table 2:** Characteristics related to nutrition and morbidity among children under-five years attending health facilities in Kombolcha town, Northeast Ethiopia (n=395).

| Variables | Category | Frequencies | Percent |
| --- | --- | --- | --- |
| History of malaria in children | Yes | 14 | 3.5 |
|  | No | 381 | 96.5 |
| History of TB | Yes | 2 | 0.5 |
|  | No | 393 | 99.5 |
| Vitamin A supplement and deworming | Yes | 146 | 37 |
|  | No | 249 | 63 |
| Diarrhea in children | Yes | 171 | 43.3 |
|  | No | 224 | 56.7 |
| Exclusive breast feeding | Yes | 173 | 43.7 |
|  | No | 222 | 56.2 |
| Dietary diversity score | Low | 139 | 35.2 |
|  | Adequate | 256 | 64.8 |

**Abbreviation:** TB, Tuberculosis

### Anemia among children

The mean hemoglobin value of children was 11.4 g/dl (SD ±18.3). From all children 206 (52.2%; 95% CI 46.8-57.0) were anemic, of which 114 (55.3%) were males (Fig 1).

**Fig 1:**
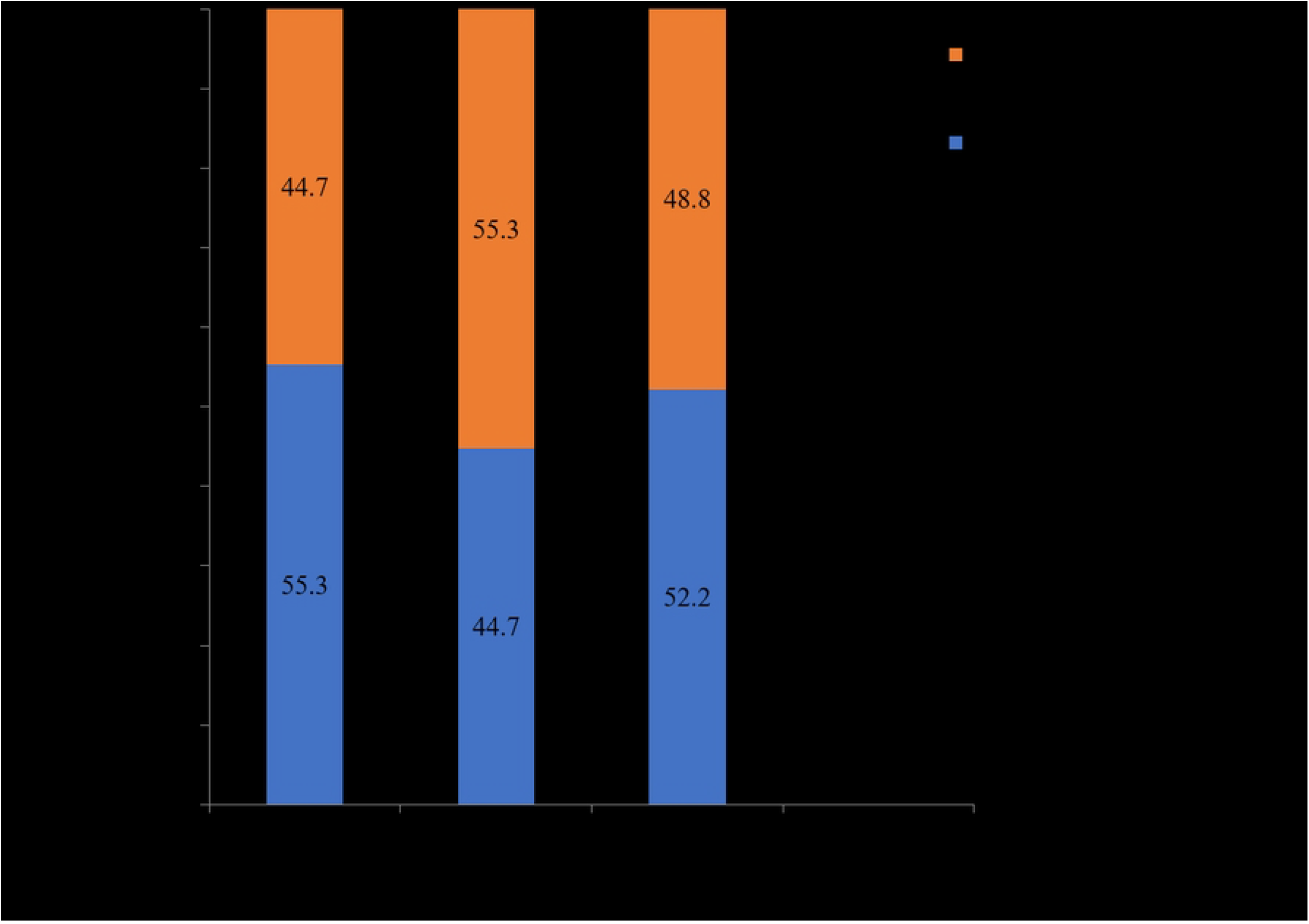
Proportion of anemia aggregated by child sex.

### Factors associated with anemia

Children in the age group of 6-11 months were six times more likely to have anemia as compared to 48-59 months (AOR: 6.23, 95% CI: 2.44, 15.95). Similarly, children 12-23 months of age were 3.74 times as likely to be anemic than 48-59 months children (AOR: 3.74, 95% CI: 1.63, 8.60).

Children from mothers of at least 30 years old were 63% less likely to be anemic as compared to children from younger mothers (AOR: 0.37, 95% CI: 0.18, 0.77). Children who exclusively breast fed up to six months were 73% less likely to be anemic compared to children who were not exclusively breast fed (AOR: 0.27, 95% CI: 0.16, 0.45). Children having low dietary diversity score were 2.6 times more likely to have anemia than those having adequate DDS (AOR: 2.61, 95% CI: 1.55, 4.38). Children who have history of diarrhea in the past 2 months prior to the data collection were nearly 2 times more likely to be anemic than children who do not have history of diarrhea (AOR: 1.87, 95% CI: 1.12, 3.12). Children from households with Family monthly income of <1,500 ETB were 17 times more likely to have anemia as compared to those who had family income of >4,500 ETB (AOR: 16.97, 95%CI: 4.95, 58.20) (Table 3).

**Table 3:** Factors associated with anemia among children 6-59 months in Kombolcha town, Northeast Ethiopia (n=395).

| Factors |  | Anemia |  | COR<br>(95% CI) | AOR<br>(95% CI) | P-value |
| --- | --- | --- | --- | --- | --- | --- |
|  |  | Yes | No |  |  |  |
| Age of Children (in months) | 6-11 | 51 | 21 | 6.38 (2.9, 13.7) | 6.23 (2.44, 15.95) | <0.001 |
|  | 12-23 | 76 | 52 | 3.84 (1.9, 7.5) | 3.74 (1.63, 8.60) | 0.002 |
|  | 24-35 | 36 | 44 | 2.14 (1.04, 4.4) | 2.31 (0.94, 5.68) | 0.067 |
|  | 36-47 | 27 | 30 | 2.36(1.08, 5.1) | 2.44 (0.93, 6.44) | 0.071 |
|  | 48-59 | 16 | 42 | 1.00 | 1.00 |  |
| Residence | Urban | 159 | 171 | 1.00 | 1.00 |  |
|  | Rural | 47 | 18 | 2.81 (1.57, 5.04) | 1.89 (0.85, 4.20) | 0.119 |
| Maternal age (in years) | <30 | 182 | 146 | 1.00 | 1.00 |  |
|  | >=30 | 24 | 43 | 0.45 (0.26, 0.77) | 0.37 (0.18, 0.77) | **0.008** |
| EBF | No | 147 | 75 | 1.00 | 1.00 |  |
|  | Yes | 59 | 114 | 0.26 (0.17, 0.40) | 0.27 (0.16, 0.45) | **<0.001** |
| DDS | Low | 134 | 59 | 4.10 (2.69, 6.24) | 2.61 (1.55, 4.38) | **<0.001** |
|  | Adequate | 72 | 130 | 1.00 | 1.00 |  |
| Diarrhea in the last 2 months | No | 104 | 120 | 1.00 | 1.00 |  |
|  | Yes | 102 | 69 | 1.71 (1.14, 2.55) | 1.87 (1.12, 3.12) | **0.017** |
| Monthly income (ETB) | <1,500 | 58 | 4 | 18 (5.8, 55.4) | 16.97 (4.95, 58.20) | **<0.001** |
|  | 1,501-3,000 | 43 | 24 | 2.2 (1.1, 4.4) | 1.61 (0.67, 3.87) | 0.283 |
|  | 3,001-4,500 | 76 | 125 | 0.75 (0.4, 1.3) | 0.64 (0.33, 1.25) | 0.190 |
|  | >4,500 | 29 | 36 | 1.00 | 1.00 |  |

**Abbreviations:** AOR, Adjusted Odds Ratio; CI, Confidence Interval; COR, Crude Odds Ratio;
DDS, Dietary Diversity Score; EBF, Exclusive Breast Feeding; ETB, Ethiopian Birr

## Discussion

This study was conducted to determine the prevalence of anemia and associated factors among children 6-59 months in northeast Ethiopia. The magnitude of anemia was found to be 52.2%, which is considered severe public health problem as of the WHO severity classification of anemia that sets 40% as a threshold [24]. This finding is comparable with findings in Southern Ethiopia [25] and Brazil [26]. But it is higher than findings from Guguftu (Northeast Ethiopia) [12], Gondar (Northwest Ethiopia) [17] and Hawassa (Southern Ethiopia) [15]. This discrepancy may be justified by the fact that the area, where this study was conducted, is frequently affected by drought so that there might be food scarcity. However, this finding is lower than findings from Bedele Hospital (Southwest Ethiopia) [27], Tanzania [28], and Ghana [29]. In addition, the current prevalence of anemia is lower than the national anemia prevalence in children under-five years age [5]. This might be attributed to the fact that the majority of the participants in the current study area consume relatively high animal-based foods, which is rich in hem iron with good bioavailability.

Younger children were at a higher risk of anemia than older children. This finding is supported by studies in Guguftu (Northeast Ethiopia) [12], Gondar town (Northwest Ethiopia) [17], Adami Tullu (South Central Ethiopia) [30], Hawassa (Southern Ethiopia) [15] and Tanzania [28]. In fact, infants began to eat complementary foods after six months old. From this time onwards up to 2 years there might be difficulty in ingesting such complementary feed and may result in deficiency of nutrients leading to higher probability of anemia. In contrary, children would have ability of eating variety of foods when they become older.

Age of mother was significantly associated with anemia. Children born from older mothers were less likely to be anemic compared to children from younger mothers. This is in line with a study in Ghana [19]. As the maternal age increases, the mother probably will have more children so that a better experience on child care and feeding. The better the care and feeding practice a child get, the lesser risk of malnutrition.

Whether a child was exclusively breast fed or not also matters the occurrence of childhood anemia. Children who exclusively breastfed until six months were less likely to be anemic than their counter parts. This finding contradicted with a study in china [31]. This discrepancy may be explained by the socio-economic differences in the study participants and study design. Exclusive breastfeeding for the first six months is a highly recommended feeding practice by the WHO. It provides the optimal feeding option by preventing diarrheal diseases, respiratory tract infections and chronic diseases [32]. This may help exclusively breastfed children to be at a lower risk of anemia.

The dietary diversity score of a child was associated with anemia. Children who had low dietary diversity score were more likely to be anemic than children who had adequate dietary diversity score. This finding is similar to study done in Hohoe Municipality, Ghana [33]. However, there was no significant association between DDS and anemia among studies conducted in Menz Gera (Central Ethiopia) [34], Kilte Awulaelo (Northern Ethiopia) [20]. In fact, eating variety of foods may provide all the necessary nutrients, prevent disease and boost immunity of the child. Hence, a child with adequate dietary diversity score may have a low risk of anemia.

The occurrence of anemia was not the same among children who had diarrhea and no diarrhea. The odds of anemia among children who had diarrhea for the last two months before the date interview was almost two times higher than their counter parts. This finding contradicted with a study in University of Gondar specialized hospital (Northwest Ethiopia) [23]. This contradiction can be explained by the difference in the study participants. The current study was conducted among ambulatory patients while the study in University of Gondar specialized hospital was conducted among admitted pediatric patients. On the other hand, the University of Gondar hospital study considered acute diarrhea so that there might be a decrease in body fluid and mask the low level of hemoglobin value. Whereas, chronic diarrhea may result in depletion of important nutrients including iron due to poor absorption in the gastro-intestinal tract. But diarrhea had no association with anemia among studies conducted in Gondar (Northwest Ethiopia) [17] and Tigray (Northern Ethiopia) [35].

As family monthly income increased, the prevalence of anemia decreased markedly similar to the findings in Tigray region (Northern Ethiopa) [36], Bedele Hospital (Southwest Ethiopia) [27], Guguftu Health Center (Northeast Ethiopia) [12] and Sudan [37]. The relationship between socio-economic status and malnutrition is well established. Low socio-economic status may result in food scarcity, poor hygiene and poor child care in general. This in turn may precipitate the occurrence of malnutrition including iron deficiency anemia.

The result of this study indicated that anemia is a major public health problem among children under-five years attending health institutions of Northeast Ethiopia. According to WHO classification for persistent anemia in a population (40%) and above, this finding confirmed that anemia among under-five children was a severe public health problem. Such a high magnitude of anemia among under-five children corroborates the finding of EDHS affirming the fact that despite the implementation of micro nutrient deficiency prevention and control guideline for several years. Endorsement and implementation of the national nutrition strategy and the greater emphasis given to anemia in the newly endorsed food and nutrition policy, there remains much to be done to cure the problem anemia. The findings call for galvanizing community level intervention that really addresses key behaviors such as breast feeding and dietary behaviors.

## Limitation of the study

Anemia was measured based on only hemoglobin concentration parameter, inclusion of other measures such as serum ferritin could lead to better differentiation of cases. Being an institution-based study, the result cannot be extrapolated to the larger community. Moreover, the study was done only in one season and may not represent the magnitude for the year round.

## Conclusions

The burden of anemia among children aged 6–59 months in the study site was high and it has severe public health problem according to the WHO cut-off points. Age of child, Age of mother, family monthly income, exclusive breast-feeding status, and having low dietary diversity score is significantly associated with anemia. The health care workers are advised to educate the pregnant, lactating mothers and the whole community about good dietary practice, the benefit of exclusive breast-feeding and income-generating activities by collaborating with the finance and economy development sectors. Researchers are recommended to use additional biomarkers to differentiate the cause of anemia and its seasonality using longitudinal designs.

## Data Availability

All relevant data are within the manuscript and its Supporting Information files.

## Acknowledgments

We highly acknowledge study participants and data collectors for their keen cooperation in getting quality data.

## Supporting information

**S1 File. Ethical approval**

(PDF)

**S2 File. STROBE checklist**

(DOCX)

**S1 Dataset. Anemia and associated factors among children aged 6-59 months**.

(SAV)

